# Tocilizumab is associated with reduction in inflammation and improvement in P/F ratio in critically sick COVID19 patients

**DOI:** 10.1101/2020.10.20.20210195

**Authors:** Muhammad Asim Rana, Mubashar Hashmi, Muhammad Muneeb Ullah Saif, Muhammad Faisal Munir, Ahad Qayyum, Rizwan Pervaiz, Muhammad Mansoor Hafeez

## Abstract

**Introduction:** Coronavirus disease 2019 was initially detected in China and has been declared a global pandemic by World Health Organization on March 11, 2020. In the majority of patients, SARS□CoV□2 causes a mild to moderate illness characterized by fever and respiratory symptoms, with or without evidence of pneumonia. The recent studies suggest that anti-cytokine targeted therapies might be associated with benefit for patients with severe COVID-19 especially in improving respiratory failure. Tocilizumab, a monoclonal antibody against interleukin 6 (IL6) receptor, is associated with clinical benefit for COVID-19 patients as it inhibits IL6 and decreases inflammation.

**Methods:** As Tocilizumab has been an important part of our treatment and a strict criterion was followed to administer Tocilizumab, a retrospective study design used to assess the beneficial effects of Tocilizumab in improvement of ratio partial pressure of arterial Oxygen and fraction of inspired Oxygen (PaO_2_/FiO_2_ or P/F ratio) and C-reactive protein (CRP) in COVID19 patients has been done. 60 patients were taken for this study by using convenient sampling technique the data of demographics, laboratory results, and clinical outcomes i.e. improvement of respiratory failure depicted in the form of PF Ratio were obtained from the medical records, Statistical analysis was done with SPSS, version 21.0.

**Results:** Sixty patients (47 males and 13 females) with COVID□19 were included in this study, the mean age of patients was 53.83 (14□81) years. After administration of Tocilizumab the lab parameters were changed as CRP decreased down to .40 (9.6□73) mg/L but other parameters were not affected. The PF ratio improved in COVID-19 patients after administration of Tocilizumab the median of PF Ratio before treatment was 108 (52□362) and improved up to 128 (37□406) after Tocilizumab therapy.

**Conclusion:** In summary, Tocilizumab appears to be associated with improvement in P/F Ratio and CRP in COVID19 patients but other markers did not improve in response to Tocilizumab therapy in severely ill COVID-19 patients.

## Introduction

Coronavirus disease 2019 (COVID19), caused by severe acute respiratory syndrome coronavirus 2 (SARS-CoV2), was initially detected in China and was later declared a global pandemic by World Health Organization on March 11, 2020 (1).

The clinical spectrum of this illness ranges from asymptomatic infection to acute respiratory distress syndrome (ARDS) (2). In the majority of patients, SARS□CoV□2 causes a mild to moderate illness characterized by fever and respiratory symptoms, with or without evidence of pneumonia.(3). However, up to 10% of patients with COVID 19 may develop severe pneumonia with hypoxia, acute respiratory distress syndrome, and multi organ failure. Such patients may require admission to an intensive care unit (ICU) for critical support and mechanical ventilation. (4).

Patients with severe COVID 19 quickly progressed to acute respiratory failure, pulmonary edema and acute respiratory distress syndrome (ARDS) (5). ARDS is the leading cause of mortality in COVID 19 patients (4). This does not occur due to uncontrolled viral replication alone but also because of an uncontrolled immune reaction from the host. In existence of uncontrolled viral replication, the presence of an increased number of damaged epithelial cells and cell debris activate a massive cytokine release also known as cytokine storm (6).

Elevated pro-inflammatory cytokines and inflammatory biomarkers such as interleukin 6, C-reactive protein (CRP), and ferritin have been shown to be higher in patients with severe COVID-19 and predictors of mortality (7, 8).

In one study, the level of interleukin 6 was noted to have increased tenfold in critically ill COVID 19 patients who required mechanical ventilation or vasopressor support in comparison to those with milder disease and interleukin 6 level correlated with the detection of SARS-CoV-2 RNA (9). These findings suggest that anti-cytokine targeted therapies may be of benefit for patients with severe COVID19. Tocilizumab, a monoclonal antibody against interleukin 6 receptor, was previously approved by Food and Drug Administration for the treatment of cytokine release syndrome (10) and may provide clinical benefit for selected COVID-19 patients with high inflammatory biomarkers.

## METHODOLOGY

The patients infected with COVID19, who were treated with Tocilizumab from May 1^st^ to 5^th^ July 2020 at Bahria international Hospital Lahore, were recruited in this retrospective study. All patient identities were kept anonymous. The study was approved by the ethical committee of Bahria International Hospital Lahore. The data utilized, including demographics, laboratory results, and clinical outcomes (PF Ratio) of the patients were obtained from medical records, based on diagnosis and treatment of Pneumonia infected by COVID19 that was classified into four types: mildly ill, moderately ill, seriously ill and critically ill. The serum levels of CRP, D-Dimers and serum Ferritin level observed before and after Tocilizumab administration. CRP, an acute phase reactant reflecting the inflammatory activity, was defined as elevated when it was higher than 5.0 mg/L. The most recent CRP, D-Dimers and serum Ferritin level Tocilizumab administration was selected as the value of before Tocilizumab therapy and the changes of the value after Tocilizumab administration was observed for 10 days. The clinical outcome of the patients was evaluated within 10 days after Tocilizumab therapy. Statistical analysis was done with SPSS, version 21.0. Data were presented as median (min□max) or as the number and percentage, as appropriate. The Wilcoxon signed□rank test used to compare parameters whenever appropriate. A P□value of less than .05 was considered statistically significant.

## RESULTS

Sixty patients (47 males and 13 females) with COVID□19 were included in this study. The characteristics of patients, the use of Tocilizumab and other anti□inflammatory drugs are summarized in results. The mean age (min□max) of the patients was 53.83 (14□81) years. From a total of 60, 4(6.6%) patients were mildly ill, 9 (14.8%) patients were moderately ill, 27 (44.3%) patients were seriously ill, and 20 (32.8%) patients were critically ill.

All (100%) patients received Tocilizumab, the dose of Tocilizumab used in patients was ranged from 400 to 800 mg (4-8 mg/kg). The laboratory findings of the 60 patients before, and in the first few days summarized below.

The principal concern of this study was to see the effect of Tocilizumab with regards to improving the PF Ratio and CRP reduction in Covid-19 infected patients as mentioned in results PF ratio improved in COVID-19 patients after administration of Tocilizumab the median of PF Ratio before treatment was 108 (52□362) and improved up to 128 (37□406) after Tocilizumab therapy.

Laboratory perimeters included in this study were CRP. Serum Ferritin, D-Dimers and LDH, values of these parameters before administration of Tocilizumab were highly raised. Mean CRP (min□max) before administration of Tocilizumab was 110 (8□239) mg/L. D-Dimers before Tocilizumab were 567 (141□1620), whilst Serum Ferritin was 105 (831□1678) and similarly serum LDH was also elevated 128 (447□1485) according to the given results above CRP levels were far above the normal range in all patients before the start of Tocilizumab therapy and were rapidly ameliorated after the Tocilizumab treatment.

After administration of Tocilizumab the lab parameters were changed as CRP decreased down to 0.40 (9.6-73) mg/L but other parameters did not decrease as significantly. D-Dimers after Tocilizumab therapy was 149 (529□1600), serum Ferritin 183 (823□1637) and LHD increased to 186 (486□2717). Importantly the effect of Tocilizumab on CRP was remarkable as it decreased from 110 (8□239) mg/L to .40 (9.6□73) mg/L (P<.05), in this regard other parameters were constant or changed minimally. Although Tocilizumab has benefits in relieving inflammatory activity, for critically ill patients who received only a single dose of Tocilizumab therapy, prognosis was still unfavorable and the CRP level in the other 5 patients failed to return to normal range during the 10 days long session. In the other patients, CRP levels were in or near the normal range within 10 days. To measure the median difference of lab values and effectiveness of Tocilizumab on those parameters Wilcoxon’s Rank test used.

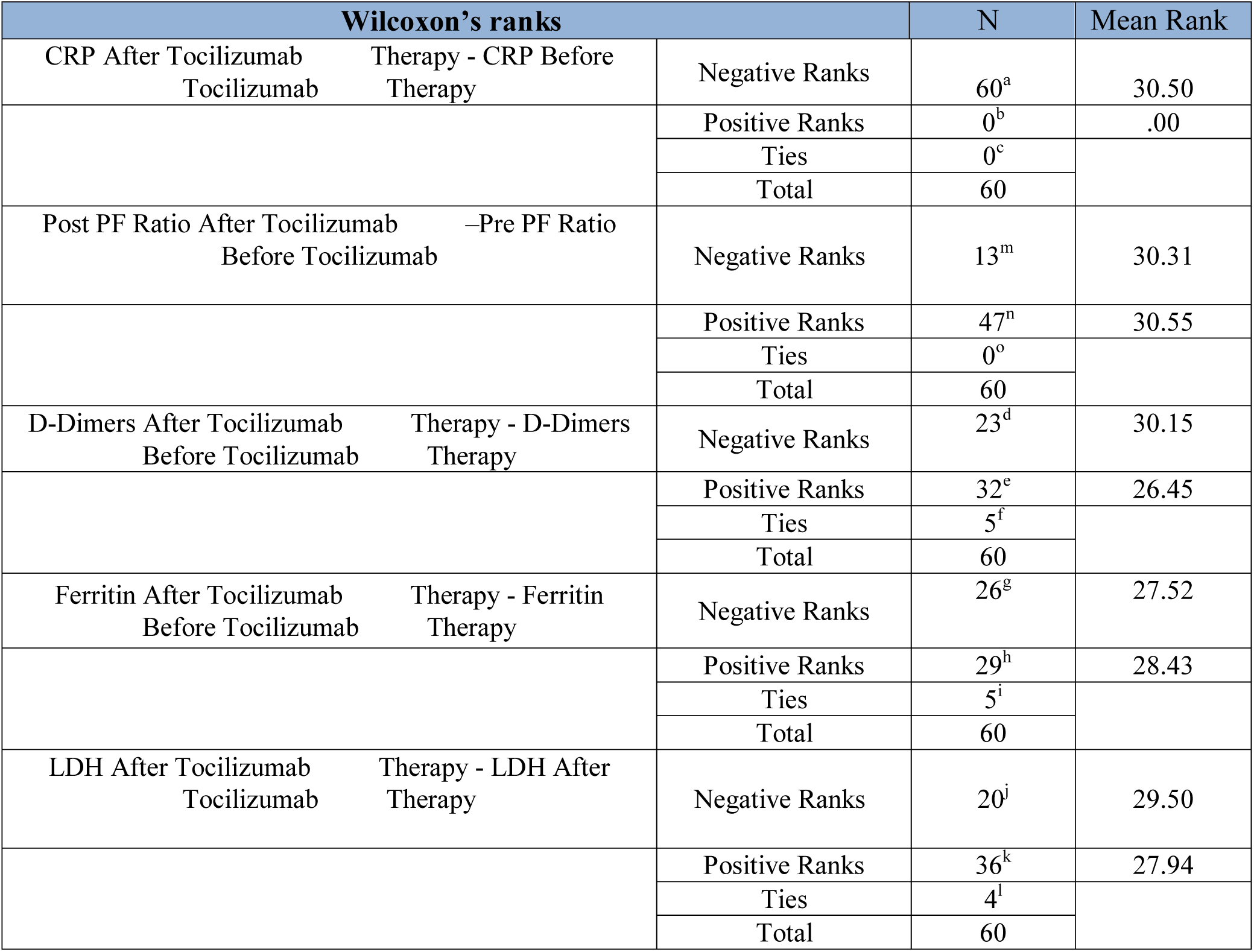

The following table elaborates on the Wilcoxon’s rank test description according to this table the 1^st^ pair describe the median CRP before and after Tocilizumab therapy in this case three ranks were seen. In these results all 60 were negative ranks, exhibiting that the median of CRP in all patients decreased significantly in posttest.

The 2^nd^ row describes the effect associated with Tocilizumab therapy in improvement of P/F Ratio according to given results there are 47 Positive Ranks and 13 negative Ranks showing that in maximum cases P/F ratio was Low before Tocilizumab administration and improved after Tocilizumab therapy, only 13 cases did not improve after Tocilizumab administration.

The 3^rd^ pair reports the D-Dimers median before and after, there were 23 negative ranks, 32 positive ranks and 5 ties mean pre and post equal. According to the given values of D-dimers only 23 ranks shows the decrease in post-test 32 positive ranks showed the increase in post-test so D-Dimers values have not decreased as significantly as CRP, after Tocilizumab therapy.

In the case of Ferritin and LDH the results were almost identical to D-Dimers. In the Ferritin row only 25 were negative ranks, 29 were positive and there were 5 ties which described that Ferritin also remained same or higher. According to last row the median LDH was very similar to Ferritin because there were 20 negative 36 positive and 4 ties which elaborated that the majority of patients LDH was high in post-test and some cases presented same value of LDH in pre and post test results.

**Table No: 2.** describes the test statistics of lab values, P/F Ratio and effects of Tocilizumab on these variables in pair of CRP and P/F Ratio before and after the P-value was .000 that was less than 0.05 that showed the beneficial effect associated with use of Tocilizumab in decreasing of CRP and improving of P/F Ratio to a remarkable level. In D-Dimers case significant value of D-dimers before and after was .522 that was greater than .05 similarly in the 3^rd^ and 4^th^ pair significant value is greater than P-value so other lab values were not decreased in response to Tocilizumab.

| Test Statistics |  |  |  |  |  |
| --- | --- | --- | --- | --- | --- |
|  | CRP after<br>Tocilizumab<br>therapy - CRP<br>before<br>Tocilizumab<br>therapy | D-dimers after<br>Tocilizumab<br>therapy - D-dimers<br>before<br>Tocilizumab<br>therapy | Ferritin after<br>Tocilizumab<br>therapy -<br>Ferritin before<br>Tocilizumab<br>therapy | LDH after<br>Tocilizumab<br>therapy - LDH<br>after<br>Tocilizumab<br>therapy | Post PF Ratio<br>after<br>Tocilizumab<br>- Pre PF ratio<br>before<br>Tocilizumab |
| Z | -6.736<sup>b</sup> | -.641<sup>c</sup> | -.663<sup>c</sup> | -1.697<sup>c</sup> | -3.835<sup>d</sup> |
| Asymp. Sig. (2-tailed) | .000 | .522 | .507 | .090 | .000 |
| a. Wilcoxon Signed Ranks Test, b. Based on positive ranks, c. Based on negative ranks, d. Based on negative ranks. |  |  |  |  |  |

## DISCUSSION

A study described the clinical characteristics and outcomes of 25 patients who received Tocilizumab therapy for severe COVID□19. Unsurprisingly, all patients were in ICU at the time of Tocilizumab initiation(11).

In our study sixty patients (78%) 47 males and (21%) 13 females with COVID19 were included. The characteristics of patients, the use of Tocilizumab and other anti-inflammatory drugs are summarized in results. The mean age (min-max) of the patients was 53.83 (14□81) years.

All (100%) patients received Tocilizumab. The dose of Tocilizumab used in patients was in the range from 400 to 800 mg. The laboratory findings of the 60 patients before, and in the first few days.

Improved P/F ratio in COVID19 patients was associated with administration of Tocilizumab the median of P/F Ratio before treatment was 108 (52□362) and improved up to 128 (37□406) after Tocilizumab therapy.

Another study narrated that at admission, 25 patients had fever, which resolved in 24 patients within 24 hours of the treatment. Furthermore, PaO_2_/FiO_2_ improved during the follow-up period (baseline, 152 ± 53; day 7, 283.73 ± 115.9; day 14, 302.2 ± 126; P < .05). Also, the CRP, ferritin, and D-dimer levels and lymphocyte count improved (12).

In our study we tried to evaluate the beneficial effects associated with use of Tocilizumab in treatment of Covid19 patients. Our findings indicated that Tocilizumab appeared to be associated with beneficial effects in decreasing the inflammatory markers significantly. In most patients, acute phase reactant levels were also decreased and the patients were achieving a relatively stable condition reflected by a decrease in CRP value after Tocilizumab administration. In this study the CRP was 110 (8-239) mg/L before Tocilizumab administration and decreased to 0.40 (9.6-73) mg/L after Tocilizumab therapy.

Similar results were shown in a study conducted in March 2020 described the effectiveness of Tocilizumab in treatment of Covid-19 Tocilizumab proved to be a significant factor in the improvement of oxygenation and decreasing CRP back to physiological levels and a later gradual decrease of IL□6 may benefit from the inhibition of inflammatory activity by Tocilizumab resulting in stabilization or improvement of clinical outcome(13).

Moreno-Pérez O described the CRP levels were far above the normal range in all patients before the start of Tocilizumab therapy, and were rapidly ameliorated after the Tocilizumab treatment. The value of CRP Post-Tocilizumab therapy was significantly decreased compared with Pre-Tocilizumab therapy, which dropped from 126.9 (10.7□257.9) to 11.2 (0.02□113.7) mg/L (P < .01). (14)

## CONCLUSION

In summary, Tocilizumab was associated with improvement in P/F Ratio and CRP significantly in COVID19 patients but there was not a noteworthy change with regards to other markers after administration of Tocilizumab in severely ill COVID19 patients. Tocilizumab seems to be an effective adjunct in treatment of severely sick COVID19 patients, which could provide a possible therapeutic strategy for this fatal disease, furthermore our result should be evaluated with caution despite the fact that we reported a good response in patients with Tocilizumab. The sample size was small and using laboratory parameters to define disease activity is challenging.

## Data Availability

All data is available with hospital IRBEC

